# Refining the impact of genetic evidence on clinical success

**DOI:** 10.1101/2023.06.23.23291765

**Authors:** Eric Vallabh Minikel, Jeffery L Painter, Coco Chengliang Dong, Matthew R. Nelson

## Abstract

The cost of drug discovery and development is driven primarily by failure, with just ∼10% of clinical programs eventually receiving approval. We previously estimated that human genetic evidence doubles the success rate from clinical development to approval. In this study we leverage the growth in genetic evidence over the past decade to better understand the characteristics that distinguish clinical success and failure. We estimate the probability of success for drug mechanisms with genetic support is 2.6 times greater than those without. This relative success varies among therapy areas and development phases, and improves with increasing confidence in the causal gene, but is largely unaffected by genetic effect size, minor allele frequency, or year of discovery. These results suggest we are far from reaching peak genetic insights to aid the discovery of targets for more effective drugs.

---

The cost of drug discovery and development is driven primarily by failure^1^, with just ∼10% of clinical programs eventually receiving approval^2–4^. We previously estimated that human genetic evidence doubles the success rate from clinical development to approval^5^. In this study we leverage the growth in genetic evidence over the past decade to better understand the characteristics that distinguish clinical success and failure. We estimate the probability of success for drug mechanisms with genetic support is 2.6 times greater than those without. This relative success varies among therapy areas and development phases, and improves with increasing confidence in the causal gene, but is largely unaffected by genetic effect size, minor allele frequency, or year of discovery. These results suggest we are far from reaching peak genetic insights to aid the discovery of targets for more effective drugs.

Human genetics is one of the only forms of scientific evidence that can demonstrate the causal role of genes in human disease. It provides a crucial tool for identifying and prioritizing potential drug targets, providing insights into the expected effect (or lack thereof^6^) of pharmacological engagement, dose-response relationships^7–10^, and safety risks^6,11–13^. Nonetheless, many questions remain about the application of human genetics in drug discovery. Genome-wide association studies (GWAS) of common, complex traits, including many diseases, generally identify variants of small effect. This contributed to early skepticism of the value of GWAS^14^. Anecdotally, such variants can point to highly successful drug targets^7–9^, and yet, genetic support from GWAS is somewhat less predictive of drug target advancement than support from Mendelian disease^5,15^.

In this paper we investigate several open questions regarding the use of genetic evidence for prioritizing drug discovery. We explore the characteristics of genetic associations that are more likely to differentiate successful from unsuccessful drug mechanisms, exploring how they differ across therapy areas and among discovery and development phases. We also investigate how close we may be to saturating the insights we can gain from genetic studies for drug discovery and how much of the genetically-supported drug discovery space remains clinically unexplored.

To characterize the drug development pipeline, we filtered Citeline Pharmaprojects for monotherapy programs added since 2000 annotated with a highest phase reached and assigned both a human gene target (usually the gene encoding the drug target protein) and an indication defined in Medical Subject Headings (MeSH) ontology. This resulted in 29,476 target-indication (T-I) pairs for analysis (Extended Data Fig. 1A). Multiple sources of human genetic associations totaled 81,939 unique gene-trait (G-T) pairs, with traits also mapped to MeSH terms. Intersection these datasets yielded an overlap of 2,166 T-I and G-T pairs (7.3%) where the indication and the trait MeSH terms had a similarity ≥0.8; we defined these T-I pairs as possessing genetic support (Extended Data Fig. 1B, 2A, see Methods). The probability of having genetic support, or P(G), was higher for launched T-I pairs than those in historical or active clinical development (Figure 1A). In each phase, P(G) was higher than previously reported^5,15^, owing, as expected^15,16^, more to new G-T discoveries than to changes in drug pipeline composition (Extended Data Fig. 3A-F). For ensuing analyses, we considered both historical and active programs. We defined success at each phase as a T-I pair transitioning to the next development phase (e.g. from phase I to II), and we also considered overall success — advancing from phase I to a launched drug. We defined relative success (RS) as the ratio of the probability of success, P(S), with genetic support to the probability of success without genetic support (see Methods). We tested the sensitivity of RS to various characteristics of genetic evidence. RS was sensitive to the indication-trait similarity threshold (Extended Data Fig. 2A), which we set to 0.8 for all analyses herein. RS was >2 for all sources of human genetic evidence examined (Figure 1B). RS was highest for OMIM (RS = 3.7), in agreement with prior reports^5,15^; this was not the result of a higher success rate for orphan drug programs (Extended Data Fig. 2B), a designation commonly acquired for rare diseases. Rather, it may owe partly to the difference in confidence in causal gene assignment between Mendelian conditions and GWAS, supported by the observation that the RS for Open Targets Genetics (OTG) associations was sensitive to the confidence in variant-to-gene mapping as reflected in the minimum share of locus-to-gene (L2G) score (Fig. 1C). The differences common and rare disease programs face in regulatory and reimbursement environments^4^ and differing proportions of drug modalities^9^ likely contribute as well. OMIM and GWAS support were synergistic with one another (Fig. S2B). Somatic evidence from IntOGen had an RS of 2.3 in oncology (Extended Data Fig. 2C), similar to GWAS, but analyses below are limited to germline genetic evidence unless otherwise noted.

**Figure 1.**
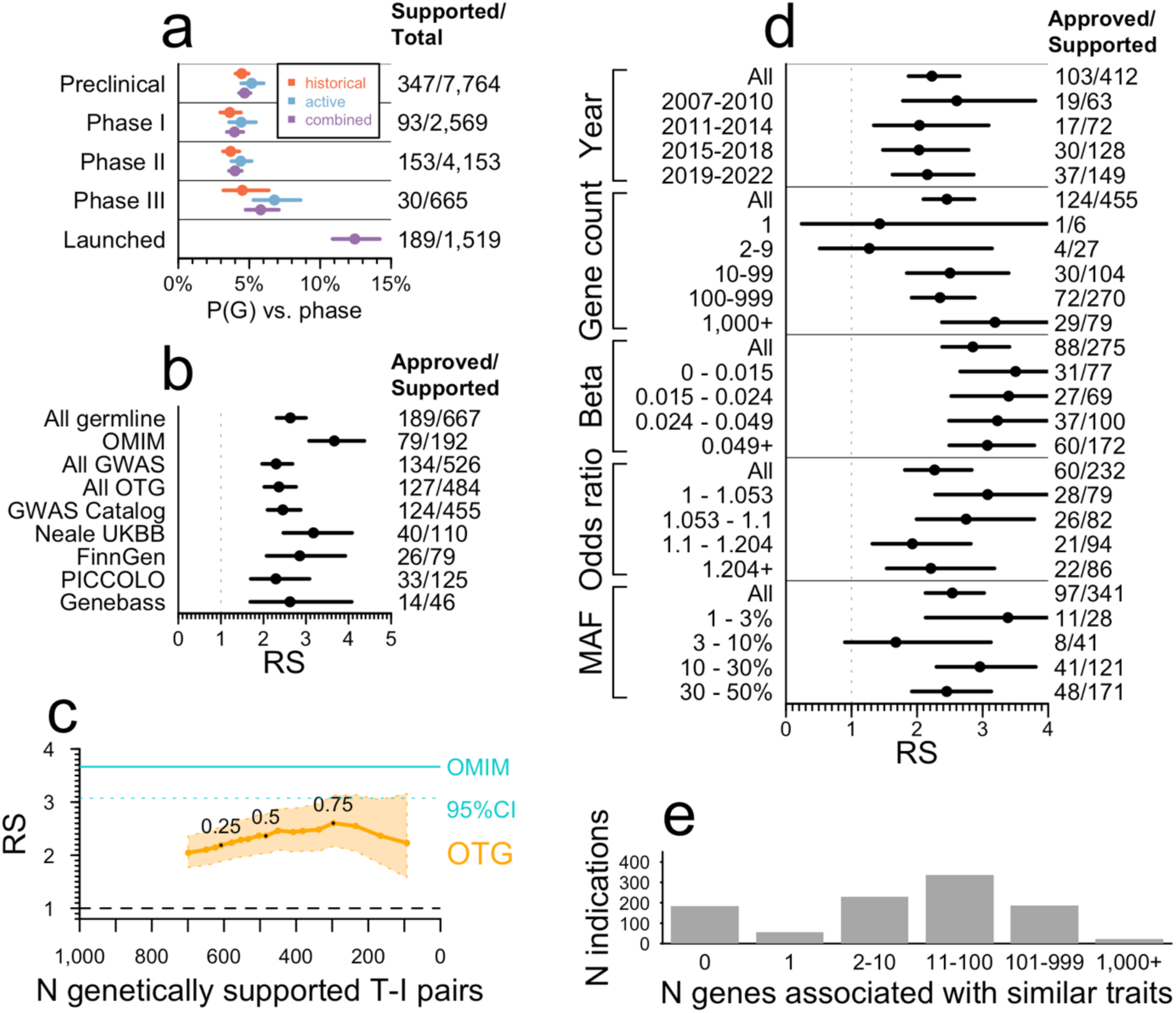
Impact of genetic evidence characteristics on relative success. **A)** Proportion of target-indication (T-I) pairs with genetic support, P(G), as a function of highest phase reached. Bars are Wilson 95% confidence intervals. **B)** Sensitivity of relative success (RS) from phase I– launch of T-I pairs with genetic evidence to source of human genetic association. GWAS Catalog, Neale UKBB, and FinnGen are subsets of Open Targets Genetics (OTG). Bars are Katz 95% confidence intervals. **C)** Sensitivity of RS to locus-to-gene (L2G) share threshold among OTG genome-wide association study (GWAS) significant associations. The minimum L2G share required for inclusion in the dataset is varied from 0.1 to 1.0 in increments of 0.05 (labels) while RS (y axis) is plotted against the number of clinical (phase I+) programs considered to have genetic support from OTG (x axis). Shaded areas are Katz 95% confidence intervals. **D)** Sensitivity of RS for OTG GWAS-supported T-I pairs to binned variables: i) year in which a T-I pair first acquired human genetic support from GWAS, excluding replications and excluding T-I pairs otherwise supported by OMIM, ii) number of genes exhibiting genetic association to the same trait, iii) quartile of effect size (beta) for quantitative traits, iv) quartile of effect size (odds ratio, OR) for case/control traits standardized to be >1 (i.e., 1/OR if <1), and v) order of magnitude of minor allele frequency bins. Bars are Katz 95% confidence intervals. **E)** Count of indications ever in development in Pharmaprojects (y axis) by the number of genes associated with traits similar to those indications (x axis). See Figure S1 for the same analyses restricted to drugs with a single known target.

As sample sizes grow ever larger with a corresponding increase in the number of unique G-T associations, some expect^17^ the value of GWAS genetic findings to become less useful for the purpose of drug target selection. We explored this in several ways. We investigated the year that genetic support for a T-I pair was first discovered, under the expectation that more common and larger effects are discovered earlier. Although there was a slightly higher RS for discoveries from 2007-2010 that was largely driven by early lipid and cardiovascular-related associations, the effect of year was overall non-significant (*P* = 0.46, Fig. 1D). Results were similar when replicate associations or OMIM discoveries were included (Extended Data Fig. 2D-F). We next divided up GWAS-supported drug programs by the number of unique traits associated to each gene. RS nominally increased with the number of associated genes, by 0.048 per gene (*P* = 0.024, Fig. 1D). This is unlikely due to successful genetically-supported programs inspiring other programs, as most genetic support was discovered retrospectively (Extended Data Fig. 2G); the few examples of drug programs prospectively motivated by genetic evidence were primarily for Mendelian diseases^9^. There were no statistically significant associations with estimated effect sizes (*P* = 0.90 and 0.57, for quantitative and binary traits, respectively; Fig. 1D, Extended Data Fig. 2H) nor minor allele frequency (*P* = 0.026, Fig. 1D). That ever larger GWAS can continue to uncover support for successful targets is also illustrated by two recent large GWAS in type 2 diabetes (T2D)^18,19^ (Extended Data Fig. 4). When these GWAS quadrupled the number of T2D associated genes from 217 to 862, new genetic support was identified for 7 of 95 mechanisms in clinical development while the number supported increased from 5 to 7 out of 12 launched drug mechanisms.

Previously^5^, we observed significant heterogeneity amongst therapy areas in the fraction of approved drug mechanisms with genetic support, but did not investigate the impact on probability of success^5^. Here, our estimates of RS from phase I to launch showed significant heterogeneity (*P* < 1.0e-15), with nearly all therapy areas having estimates greater than one, 11 of 17 were >2, and hematology, metabolic, respiratory, and endocrine >3 (Fig. 2A-E). In most therapy areas, the impact of genetic evidence was most pronounced in phases II and III and least impactful in phase I, corresponding to capacity to demonstrate clinical efficacy in later development phases. Accordingly, therapy areas differed in P(G) and in whether P(G) increased throughout clinical development or only at launch (Extended Data Fig. 5); data source and other properties of genetic evidence including year of discovery and effect size also differed (Extended Data Fig. 6). We also found that genetic evidence differentiated likelihood to progress from preclinical to clinical development for metabolic diseases (RS = 1.38, 95% CI = 1.25 – 1.54), that may reflect preclinical models that are more predictive of clinical outcomes. Probability of genetic support by therapy area was correlated with probability of success, or P(S) (ρ = 0.59, *P* = 0.013) and with RS (ρ = 0.72, *P* = 0.0011; Extended Data Fig. 7), which led us to explore how the sheer quantity of genetic evidence available within therapy areas (Fig. 2F, Extended Data Fig. 8A) may influence this. We found that therapy areas with more possible gene-indication (G-I) pairs supported by genetic evidence had significantly higher RS (ρ = 0.71, *P* = 0.0010, Fig. 2G), although respiratory and endocrine were notable outliers with high RS despite fewer associations.

**Figure 2.**
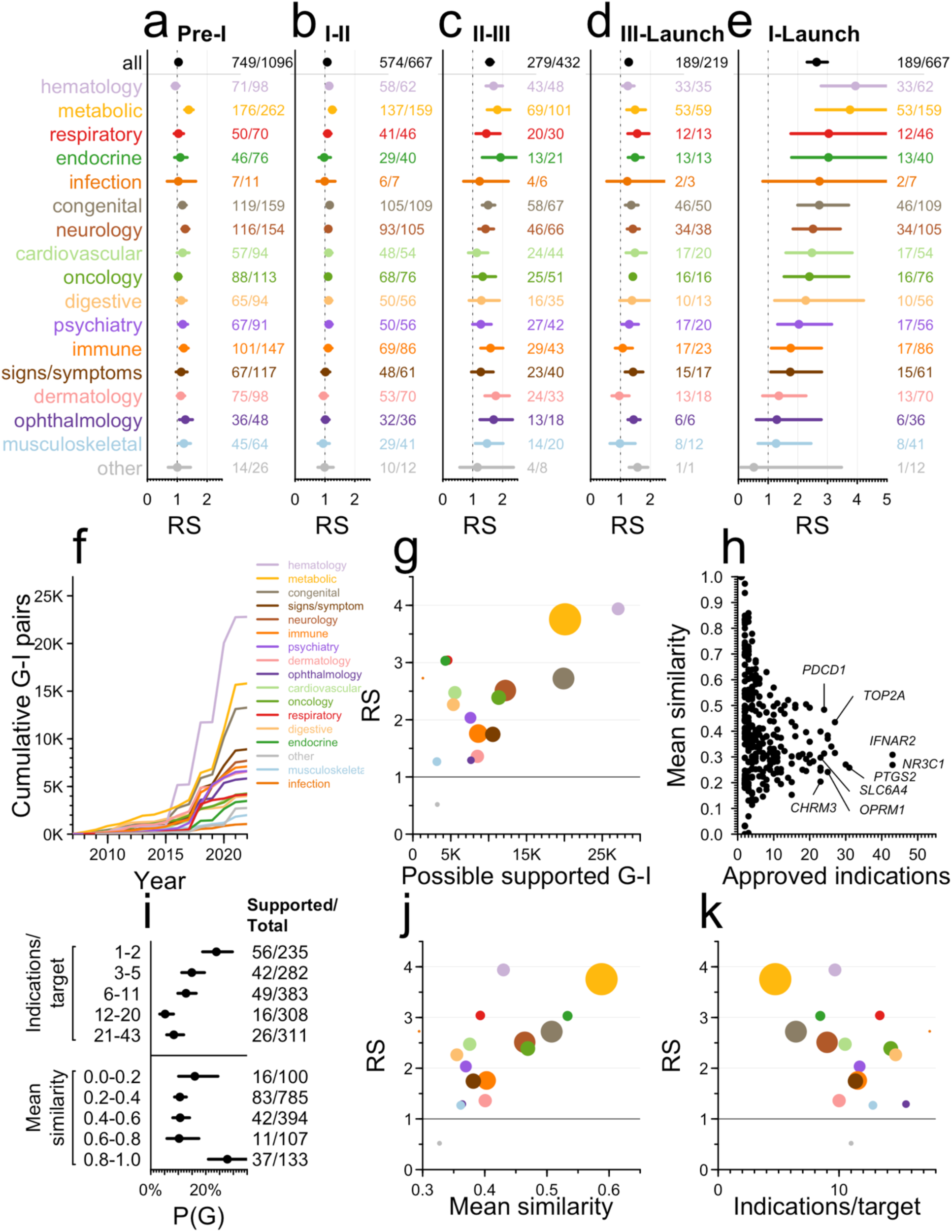
Differences in relative success between therapy areas and the number and diversity of indications per target. **A-E)** RS by therapy area and phase transition. Bars are 95% confidence intervals. **F**) Cumulative number of possible genetically supported gene-indication (G-I) pairs in each therapy (y axis) as genetic discoveries have accrued over time (x axis). **G**) RS (y axis) by number of possible supported G-I pairs (x axis) across therapy areas, dots colored as in panels A-E and sized according to number of genetically supported T-I pairs in at least phase I. **H)** Number of approved indications vs. similarity of those indications, by approved drug target. **I)** Proportion of approved target-indication pairs with genetic support, P(G), binned by quintile of the number of approved indications per target (top panel) or by mean similarity among approved indications (bottom panel). Targets with exactly 1 approved indication (6.2% of launched T-I pairs) are considered to have mean similarity of 1.0. Bars are Wilson 95% confidence intervals. **J)** RS (y axis) vs. mean similarity among approved indications per target (x axis) by therapy area. **K)** RS (y axis) vs. mean count of approved indications per target (x axis). See Figure S2 for the same analyses restricted to drugs with a single known target.

We hypothesized that genetic support might be most pronounced for drug mechanisms with disease-modifying effects, as opposed to those that manage symptoms, and that the proportion of such drugs differ by therapy area^20,21^. We were unable to find data with these descriptions available for a sufficient number of drug mechanisms to analyze, but we reasoned that targets of disease-modifying drugs are more likely to be specific to a disease, whereas targets of symptom-managing drugs are more likely to be applied across many indications. We therefore examined the number and diversity of all-time launched indications per target. Launched T-I pairs are heavily skewed towards a few targets (Fig. 2H). Of 450 launched targets, the 42 with ≥10 launched indications comprise 713 (39%) of 1,806 launched T-I pairs (Fig. 2H). Many of these are used across diverse indications for management of symptoms such as inflammatory and immune responses (*NR3C1*, *IFNAR2*), pain (*PTGS2*, *OPRM1*), mood (*SLC6A4*), or parasympathetic response (*CHRM3*). The count of launched indications was inversely correlated with the mean similarity of those indications (ρ = −0.72, *P* = 4.4e-84; Fig. 2H). Among T-I pairs, the probability of having genetic support increased as the number of approved indications decreased (*P* = 6.3e-7) and as the similarity of a target’s approved indications increased (*P* =1.8e-5, Fig. 2I). We observed a corresponding impact on RS, increasing in therapy areas where the similarity among approved indications increased, and decreasing with increasing indications per target (ρ = 0.74, *P* = 0.0010, and ρ = −0.62, *P* = 0.0080, respectively, Fig. 2J-K).

Only 4.8% (284/5,968) of T-I pairs active in phase I-III possess human germline genetic support (Figure 1A), similar to T-I pairs no longer in development (4.2%, 560/13,355), a difference that was not statistically significant (*P* = 0.080). We estimated (see Methods) that only 1.1% of all genetically supported G-I relationships have been explored clinically (Fig. 3A), or 2.1% when restricting to the most similar indication. Given that the vast majority of proteins are classically “undruggable”, we explored the proportion of genetically supported G-I pairs that had been developed to at least phase I, as a function of therapy area across several classes of tractability and relevant protein families^22^ (Fig. 3A). Within therapy areas, oncology kinases with germline evidence were the most saturated: 109 of 250 (44%) of all genetically supported G-I pairs had reached at least phase I; GPCRs for psychiatric indications were also notable (14/53, 26%). Grouping by target rather than G-I pair, 3.6% of genetically supported targets have been pursued for any genetically supported indication (Extended Data Figure 8). Of possible genetically supported G-I pairs, most (68%) arose from OTG associations, mostly within the past 5 years (Fig. 2F). Such low utilization is partly due to recent emergence of most genetic evidence (Extended Data Fig. 2F-G, 7A), since drug programs prospectively supported by human genetics have had a mean lag time from genetic association of 13 years to first trial^21^ and 21 years to approval^9^. Because some types of targets may be more readily tractable by antagonists than agonists, we also grouped by target and examined human genetic evidence by direction of effect for tumor suppressors versus oncogenes (Fig. 3B), identifying a few substrata for which a majority of genetically supported targets had been pursued to at least phase I for at least one genetically supported indication. Oncogene kinases received the most attention, with 19/25 (76%) reaching phase I.

**Figure 3.**
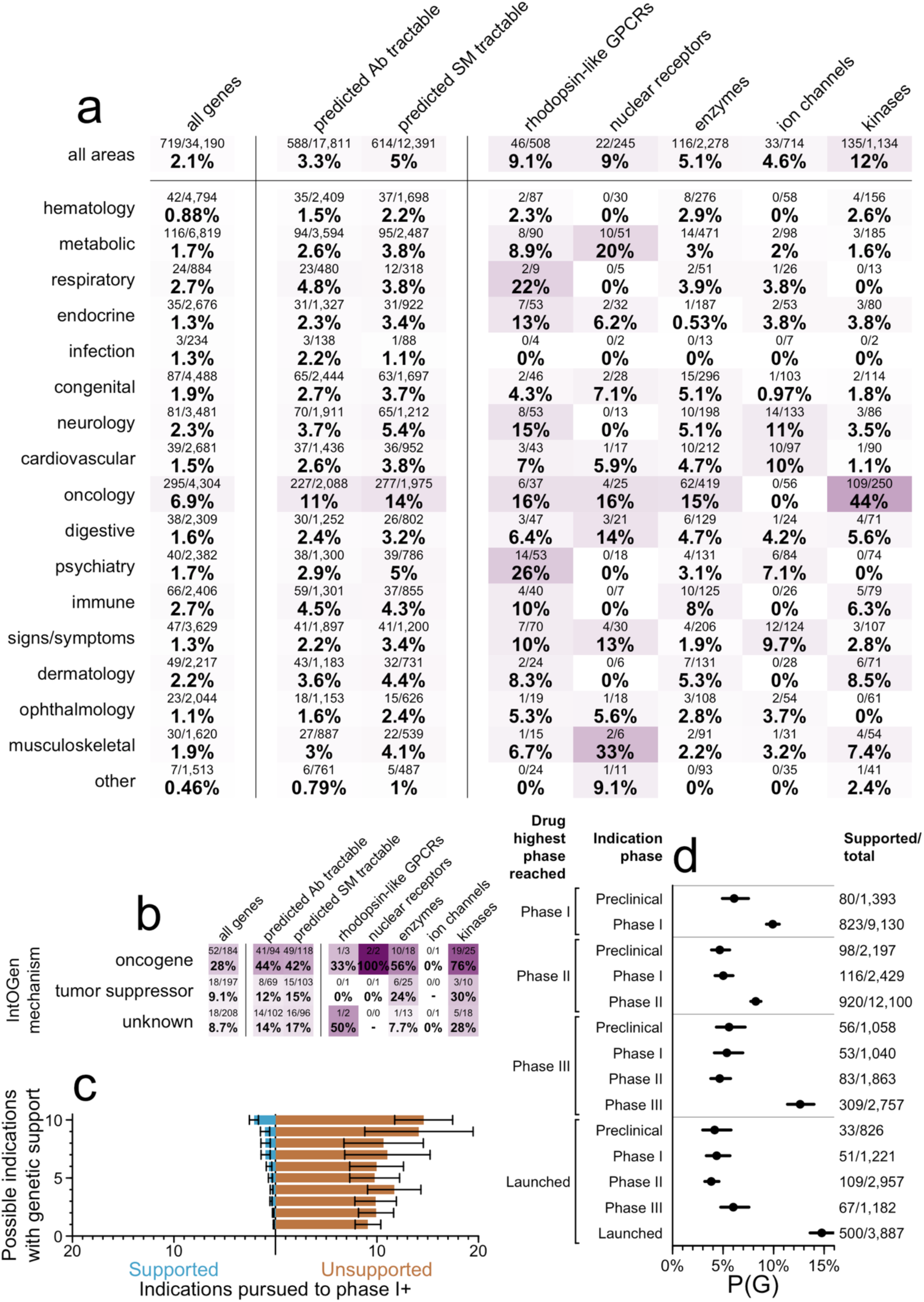
Clinical investigation of drug mechanisms with genetic evidence. **A)** Heatmap of proportion of genetically supported T-I pairs that have been developed to at least phase I, by therapy area (y axis) and gene list (x axis). **B)** As panel A, but for genetic support from IntOGen rather than germline sources and grouped by the direction of effect of the gene according to IntOGen (y axis), and also grouped by target rather than T-I pair. Thus, the denominator for each cell is the number of targets with at least one genetically supported indication, and each target counts towards the numerator if at least one genetically supported indication has reached phase I. **C)** Of targets that have both reached phase I for any indication, and have at least one genetically supported indication, the mean count (x axis) of genetically supported (left) and unsupported (right) indications pursued, binned by the number of possible genetically supported indications (y axis). Bars are Wilson 95% confidence intervals. **D)** Proportion of D-I pairs with genetic support, P(G) (x axis), as a function of each D-I pair’s phase reached (inner y axis grouping) and the drug’s highest phase reached for any indication (outer y axis grouping). Bars are Wilson 95% confidence intervals. See Figure S3 for the same analyses restricted to drugs with a single known target.

To focus on demonstrably druggable proteins, we further restricted the analysis to targets with both i) any program reaching phase I, and ii) ≥1 genetically supported indication. Out of 1,147 qualifying targets, only 373 (33%) had been pursued for one or more supported indications (Fig. 3C), and most (307, 27%) of these targets were pursued both for indications with and without genetic support. Overall, an overwhelming majority of development effort has been for unsupported indications, at a 17:1 ratio. Within this subset of targets, we asked whether genetic support was predictive of which indications would advance the furthest. Grouping active and historical programs by D-I pair, we found that the odds of advancing to a later stage in the pipeline is 82% higher for indications with genetic support (*P* = 8.6e-73, Fig. 3D).

While there have been anecdotes such as *HMGCR* to argue that genetic effect size may not matter in prioritizing drug targets, here we provide systematic evidence that small effect size, recent year of discovery, increasing number of genes identified, or higher associated allele frequency do not diminish the value of GWAS evidence to differentiate clinical success rates. One reason for this is likely because genetic effect size on a phenotype rarely accounts for the magnitude of genetic effect on gene expression, protein function, or some other molecular intermediate. In some circumstances, genetic effect sizes can yield insights into anticipated drug effects. This is best Illustrated for cardiovascular disease therapies, where genetic effects on cholesterol and disease risk and treatment outcomes are correlated^23^. A limitation is that, other than Genebass, we did not include whole exome or whole genome sequencing association studies, which may be more likely to pinpoint causal variants. Moreover, all of our analyses are naïve to direction of genetic effect (gain versus loss of gene function) as this is unknown or unannotated in most datasets utilized here.

Our results argue for continuing investment to expand GWAS-like evidence, particularly for many complex diseases with treatment options that fail to modify disease. Although genetic evidence has value across most therapy areas, its benefit is more pronounced in some areas than others. Furthermore, it is possible that the therapy areas where genetic evidence had a lower impact have seen more focus on symptom management. If so, we would predict that for drugs aimed at disease modification, human genetics should ultimately prove highly valuable across therapy areas.

The focus of this work has been on the relative success of drug programs with and without genetic evidence, limited to drug mechanisms that have entered clinical development. This metric does not address the probability that a gene associated with a disease, if targeted, will yield a successful drug. At the early stage of target selection, is evidence of a large loss of function effect in one gene usually a better choice than a small non-coding SNP effect on the same phenotype in another? We explored this question for T2D studies referenced above. Of the 7 targets of launched drugs with genetic evidence, 4 had Mendelian evidence (in addition to pre-2020 GWAS evidence), out of a total of 19 Mendelian genes related to T2D (21%). 1 launched T2D target had only GWAS (and no Mendelian) evidence among 217 GWAS associated genes prior to 2020 (0.46%), while 2 launched targets were among 645 new GWAS associations since 2020 (0.31%). At least in this example, the “yield” of genetic evidence for successful drug mechanisms was greatest for genes with Mendelian effects, but similar between earlier and later GWAS. Clearly, just because genetic associations differentiate clinical stage drug targets from launched ones does not mean that a large fraction of associations will be fruitful. Moreover, genetically supported targets may be more likely to require upregulation, to be druggable only by more challenging modalities^4,9^, or to enjoy narrower use across indications. More work is required to better understand the challenges of target identification and prioritization given the genetic evidence precondition.

The utility of human genetic evidence in drug discovery has had firm theoretical and empirical footing for several years^5,7,15^. If the benefit of this evidence were canceled out by competitive crowding^24^, then currently active clinical phases should have higher rates of genetic support than their corresponding historical phases, and might look similar to, or even higher than, approved pairs. Instead, we find that active programs possess genetic support only slightly more often than historical programs and remain less enriched for genetic support than approved drugs. Meanwhile, only a tiny fraction of classically druggable genetically supported G-I pairs have been pursued even among targets with clinical development reported. Human genetics thus represents a growing opportunity for novel target selection and improving indication selection for existing drugs and drug candidates. Increasing emphasis on drug mechanisms with supporting genetic evidence is expected to increase success rates and lower the cost of drug discovery and development.

## Methods

### Definition of metrics

Except where otherwise noted, we define genetic support of a drug mechanism (i.e. a target-indication or T-I pair) as a genetic association mapped to the corresponding target gene for a trait that is ≥0.8 similar to the indication (see MeSH term similarity below). We defined the probability of <u>g</u>enetic support, or P(G), as the proportion of drug mechanisms satisfying the above definition of genetic support. Probability of <u>s</u>uccess, or P(S), is the proportion of programs in one phase that advance to a subsequent phase (for instance, phase I to phase II). Overall P(S) from phase I to launched is the product of P(S) at each individual phase. Relative success, or RS, is the ratio of P(S) for programs with genetic support to P(S) for programs lacking genetic support, which is equivalent to a relative risk or risk ratio. Thus, if N denotes the total number of programs that have reached the reference phase, and X denotes the number of those that advance to a later phase of interest, and the subscripts G and !G indicate the presence or absence of genetic support, then P(G) = N_G_ / (N_G_ + N_!G_); P(S) = (X_G_ + X_!G_) / (N_G_ + N_!G_); RS = (X_G_/N_G_)/(X_!G_/N_!G_). RS from phase I to launched is the product of RS at each individual phase. The count of “programs” for X and N is target-indication (T-I) pairs throughout, except for Figure 3D, which uses drug-indication pairs (D-I) in order to specifically interrogate P(G) where the same drug has been developed for different indications. For clarity, we note that where other recent studies^22,25^ have examined the fold enrichment and overlap between genes with a human genetic support and genes encoding a drug target, without regards to similarity, herein all of our analyses are conditioned on the similarity between the drug’s indication and the genetically associated trait.

### Drug development pipeline

Citeline Pharmaprojects^26^ is a curated database of drug development programs including preclinical, all clinical phases, and launched drugs. It was queried via API (Dec 22, 2022) to obtain information on drugs, targets, indications, phases reached, and current development status. T-I pair was the unit of analysis throughout, except where otherwise indicated in the text (D-I pairs were examined in Figure 3D). Current development status was defined as “active” if the T-I pair had at least one drug still in active development, and “historical” if development of all drugs for the T-I pair had ceased. Targets were defined as genes; as most drugs do not directly target DNA, this usually refers to the gene encoding the protein target that is bound or modulated by the drug. We removed combination therapies, diagnostic indication, and programs with no human target or no indication assigned. For most analyses, only programs added to the database since 2000 were included, while for the count and similarity of launched indications per target, we used all launches for all time. Indications were considered to possess “genetic insight” — meaning the human genetics of this trait or similar traits have been successfully studied — if they had ≥0.8 similarity to i) an OMIM or IntOGen disease, or ii) a GWAS trait with at least 3 independently associated loci, based on lead SNP positions rounded to the nearest 1 Mb. For calculating relative success, we used the number of T-I pairs with genetic insight as the denominator. The rationale for this choice is to focus on indications where there exists the opportunity for human genetic evidence, consistent with the filter applied previously^5^. However, we observe that our findings are not especially sensitive to the presence of this filter, with RS decreasing by just 0.17 when the filter is removed (Extended Data Fig. 3G-H). Note that the criteria for determining “genetic insight” are distinct from, and much looser than, the criteria for mapping GWAS hits to genes (see locus-to-gene or L2G scores under Open Targets Genetics below). Many drugs had more than one target assigned, in which case all targets were retained for target-indication pair analyses. As a sensitivity test, running all analyses restricted to only drugs with exactly one target assigned yielded very similar results (Figures S1-S11).

### OMIM

Online Mendelian Inheritance in Man (OMIM) is a curated database of Mendelian gene-disease associations. The OMIM Gene Map (downloaded Sep 21, 2023) contained 8,671 unique gene-phenotype links. We restricted to entries with phenotype mapping code 3 (“the molecular basis for the disorder is known; a mutation has been found in the gene”), removed phenotypes with no MIM number or no gene symbol assigned, and removed duplicate combinations of gene MIM and phenotype MIM. We used regular expression matching to further filter out phenotypes containing the terms “somatic”, “susceptibility”, or “response” (drug response associations) and those flagged as questionable (“?”), or representing non-disease phenotypes (“[”). A set of OMIM phenotypes are flagged as denoting susceptibility rather than causation (“{”); this category includes low-penetrance or high allele frequency association assertions that we wished to exclude, but also germline heterozygous loss-of-function mutations in tumor suppressor genes, where the underlying mechanism of disease initiation is loss of heterozygosity, which we wished to include. We therefore also filtered out phenotypes containing “{” except for those that did contain the terms “cancer”, “neoplasm”, “tumor”, or “malignant” and did not contain the term “somatic”. Remaining entries present in OMIM as of 2021 were further evaluated for validity by two curators, and gene-disease combinations for which a disease association was deemed not to have been established were excluded from all analyses. All of the above filters left 5,670 unique gene-trait links. MeSH terms for OMIM phenotypes were then mapped using the EFO OWL database using an approach previously described^27^, with additional mappings from Orphanet, full text matches to the full MeSH vocabulary, and finally, manual curation, for a cumulative mapping rate of 93% (5,297/5,670). Because sometimes distinct phenotype MIM numbers mapped to the same MeSH term, this yielded 4,510 unique gene-MeSH links.

### Open Targets Genetics

Open Targets Genetics (OTG) is a database of GWAS hits from published studies and biobanks. OTG version 8 (October 12, 2022) variant-to-disease (V2D), locus-to-gene (L2G), variant index, and study index data were downloaded from EBI. Traits with multiple EFO IDs were excluded as these generally represent conditional, epistasis, or other complex phenotypes that would lack mappings in the MeSH vocabulary. Of the top 100 traits with the greatest number of genes mapped, we excluded 76 as having no clear disease relevance (example: “red cell distribution width”) or no obvious marginal value (example: excluded “trunk predicted mass” because “body mass index” was already included). Remaining traits were mapped to MeSH using the EFO OWL database, full text queries to the MeSH API, mappings already manually curated in PICCOLO (see below) or new manual curation. In total, 25,124/49,599 unique traits (51%) were successfully mapped to a MeSH ID. We included associations with *P* < 5e-8. OTG L2G scores used for gene mapping are based on a machine learning model trained on gold standard causal genes^28^; inputs to that model include distance, functional annotations, eQTLs, and chromatin interactions. Note that we do not utilize Mendelian randomization^29^ to map causal genes, and even gene mappings with high L2G scores are necessarily imperfect. OTG provides an L2G score for the triplet of each study or trait with each hit and each possible causal gene. We defined L2G share as the proportion of the total L2G score assigned each gene among all potentially causal genes for that trait-hit combination. In sensitivity analyses we considered L2G share thresholds from 10% to 100% (Figure 1B and Extended Data Fig. 3A), but main analyses used only genes with ≥50% L2G share (which are also the top-ranked genes for their respective associations). OTG links were parsed to determine the source of each OTG data point: the EBI GWAS catalog^30^ (N=136,503 hits with L2G share ≥0.5), Neale UK BioBank (http://www.nealelab.is/uk-biobank; N=19,139), FinnGen R6^31^ (N=2,338), or SAIGE (N=1,229).

### PICCOLO

PICCOLO^32^ is a database of GWAS hits with gene mapping based on tests for colocalization without full summary statistics by using Probabilistic Identification of Causal SNPs (PICS) and a reference dataset of SNP linkage disequilibrium values. As described^32^, gene mapping utilizes QTL data from GTEx (N=7,162) and a variety of other published sources (N=6,552). We included hits with GWAS *P* < 5e-8, and with eQTL *P* < 1e-5, and H4 ≥ 0.9, as these thresholds were determined empirically^32^ to strongly predict colocalization results.

### Genebass

Genebass^33^ is a database of genetic associations based on exome sequencing. Genebass data from 394,841 UK Biobank participants (the “500K” release) were queried using Hail (October 19, 2023). We used hits from four models: pLoF (predicted loss-of-function) or missense|LC (missense and low confidence LoF), each with SKAT or burden tests, filtering for P < 1e-5. Because the traits in Genebass are from UK Biobank, which is included in OTG, we used the OTG MeSH mappings established above.

### IntOGen

IntOGen is a database of enrichments of somatic genetic mutations within cancer types. We used the driver genes and cohort information tables (May 31, 2023). IntOGen assigns each gene a mechanism in each tumor type; occasionally a gene will be classified as a tumor suppressor in one type and an oncogene in another. We grouped by gene and assigned each gene its modal classification across cancers. MeSH mappings were curated manually.

### MeSH term similarity

MeSH terms in either Pharmaprojects or the genetic associations datasets that were Supplementary Concept Records (IDs beginning in “C”) were mapped to their respective preferred main headings (IDs beginning in “D”). A matrix of all possible combinations of drug indication MeSH IDs and genetic association MeSH IDs was constructed. MeSH term Lin and Resnik similarities were computed for each pair as described^34,35^. Similarities of −1, indicating infinite distance between two concepts, were assigned as 0. The two scores were regressed against each other across all term pairs, and the Resnik scores were adjusted by a multiplier such that both scores had a range from 0 to 1 and their regression had a slope of 1. The two scores were then averaged to obtain a combined similarity score. Similarity scores were successfully calculated for 1,006/1,013 (99.3%) of unique MeSH terms for Pharmaprojects indications, corresponding to 99.67% of Pharmaprojects T-I pairs, and for 2,260/2,262 (99.9%) unique MeSH terms for genetic associations, corresponding to >99.9% of associations.

### Therapeutic areas

MeSH terms for Pharmaprojects indications were mapped onto 16 top-level headings under the Diseases [C] and Psychiatry and Psychology [F] branches of the MeSH tree (https://meshb.nlm.nih.gov/treeView), plus an “other”. The signs/symptoms area corresponds to C23 Pathological Conditions, Signs, and Symptoms and contains entries such as inflammation and pain. Many MeSH terms map to >1 tree position; these multiples were retained and counted towards each therapy area, except for the following conditions: for terms mapped to oncology, we deleted their mappings to all other areas; and “other” was used only for terms that mapped to no other areas.

### Analysis type 2 diabetes GWAS

We included 19 genes from OMIM linked to Mendelian forms of diabetes or syndromes with diabetic features. For Vujkovic et al, 2020^18^, we considered as novel any genes with a novel nearest gene, novel coding variant, or a novel lead SNP colocalized with an eQTL with H4 ≥0.9. Non-novel nearest genes, coding variants, and colocalized lead SNPs were considered established variants. For Suzuki et al, 2023^19^, we used the available L2G scores that OTG had assigned for the same lead SNPs in previously reported GWAS for other phenotypes, yielding mapped genes with L2G share > 0.5 for 27% of loci. Genes were considered novel if absent from the Vujkovic analysis. Together, these approaches identified 217 established GWAS genes and 645 novel ones (469 from Vujkovic and 176 from Suzuki). We identified 347 unique drug targets in Pharmaprojects reported with a type 2 diabetes or diabetes mellitus indication, including 25 approved. We reviewed the list of approved drugs and eliminated those where there were questions around the relevance of the drug or target to T2D (*AKR1B1*, *AR*, *DRD1*, *HMGCR*, *IGF1R*, *LPL, SLC5A1*). Because Pharmaprojects ordinarily specifies the receptor as target for protein or peptide replacement therapies, we also remapped the minority of programs where the ligand, rather than receptor, had been listed as target (changing *INS* to *INSR*, *GCG* to *GCGR*) To assess the proportion of programs with genetic support, we first grouped by drug and selected just one target, preferring the target with the earliest genetic support (OMIM, then established GWAS, then novel GWAS, then none). Next we grouped by target and selected its highest phase reached. Finally, we grouped by highest phase reached and counted the number of unique targets.

### Universe of possible genetically supported gene-indication pairs

In all of our analyses, targets are defined as human gene symbols, but we use the term gene-indication pair (G-I) to refer to possible genes that one might attempt to target with a drug, and target-indication pair (T-I) to refer to genes that are the targets of actual drug candidates in development. To enumerate the space of possible G-I pairs, we multiplied the N=769 Pharmaprojects indications considered here by the “universe” of N=19,338 protein-coding genes, yielding a space of N= 14,870,922 possible G-I pairs. Of these, N=101,954 (0.69%) qualify as having genetic support per our criteria. A total of 16,808 T-I pairs have reached at least Phase I in an active or historical program, of which 1,155 (6.9%) are genetically supported. This represents an enrichment compared to random chance (OR = 11.0, *P* < 1.0e-15, Fisher exact test), but in absolute terms, only 1.1% of genetically supported G-I pairs have been pursued. A genetically supported G-I pair may be less likely to attract drug development interest if the indication already has many other potential targets, and/or if the indication is but the second-most similar to the gene’s associated trait. Removing associations with many GWAS hits and restricting to the single most similar indication left a space of 34,190 possible genetically supported G-I pairs, 719 (2.1%) of which had been pursued. This small percentage might yet be perceived to reflect competitive saturation, if the vast majority of indications are undevelopable and/or the vast majority of targets are undruggable. We therefore asked what proportion of genetically supported G-I pairs had been developed to at least Phase I, as a function of therapy area cross-tabulated against Open Targets predicted tractability status or membership in canonically “druggable” protein families, using families from ref. ^22^ as well as UniProt pkinfam for kinases^36^. We also grouped at the level of gene, rather than G-I pair (Extended Data Fig. 8).

### Druggability and protein families

Antibody and small molecule druggability status was taken from Open Targets^37^. For antibody tractability, Clinical Precedence, Predicted Tractable – High Confidence, and Predicted Tractable – Medium to Low Confidence were included. For small molecules, Clinical Precedence, Discovery Precedence, and Predicted Tractable were included. Protein families were from sources described previously^22^, plus the pkinfam kinase list from UniProt^36^. To make these lists non-overlapping, genes that were both kinases and also either enzymes, ion channels, or nuclear receptors were considered to be kinases only.

### Statistics

Analyses were conducted in R 4.2.0. For binomial proportions P(G) and P(S), error bars are Wilson 95% confidence intervals, except for P(S) for phase I-launch where the Wald method is used to compute the confidence intervals on the product of the individual probabilities of success at each phase. RS at each individual phase uses Wilson 95% confidence intervals, while RS for phase I–launch is defined as a product of the three phase-wise risk ratios, with Katz 95% confidence intervals. Effect of continuous variables on probability of launch were assessed using logistic regression. Differences in RS between therapy areas were tested using the Cochran-Mantel-Haenszel chi-square test (cmh.test from the R lawstat package). Pipeline progression of drug-indication pairs conditioned on the highest phase reached by a drug was modeled using an ordinal logit model (polr with Hess=TRUE from the R MASS package). Correlations across therapy areas were tested by weighted Pearson’s correlation (wtd.cor from the R weights package); to control for the amount of data available in each therapy area, the number of genetically supported T-I pairs having reached at least phase I used as the weight. Enrichments of T-I pairs in the utilization analysis were tested using Fisher’s exact test. All statistical tests were two-sided.

### Source code availability and data availability

An analytical dataset and source code are available at https://github.com/ericminikel/genetic_support/ and are sufficient to reproduce all figures and statistics herein.

## Data Availability

All source code and data produced in the present work are contained in the supplementary material an online at https://github.com/ericminikel/genetic_support/

## SUPPLEMENTARY FIGURES

**Extended Data Figure 1.**
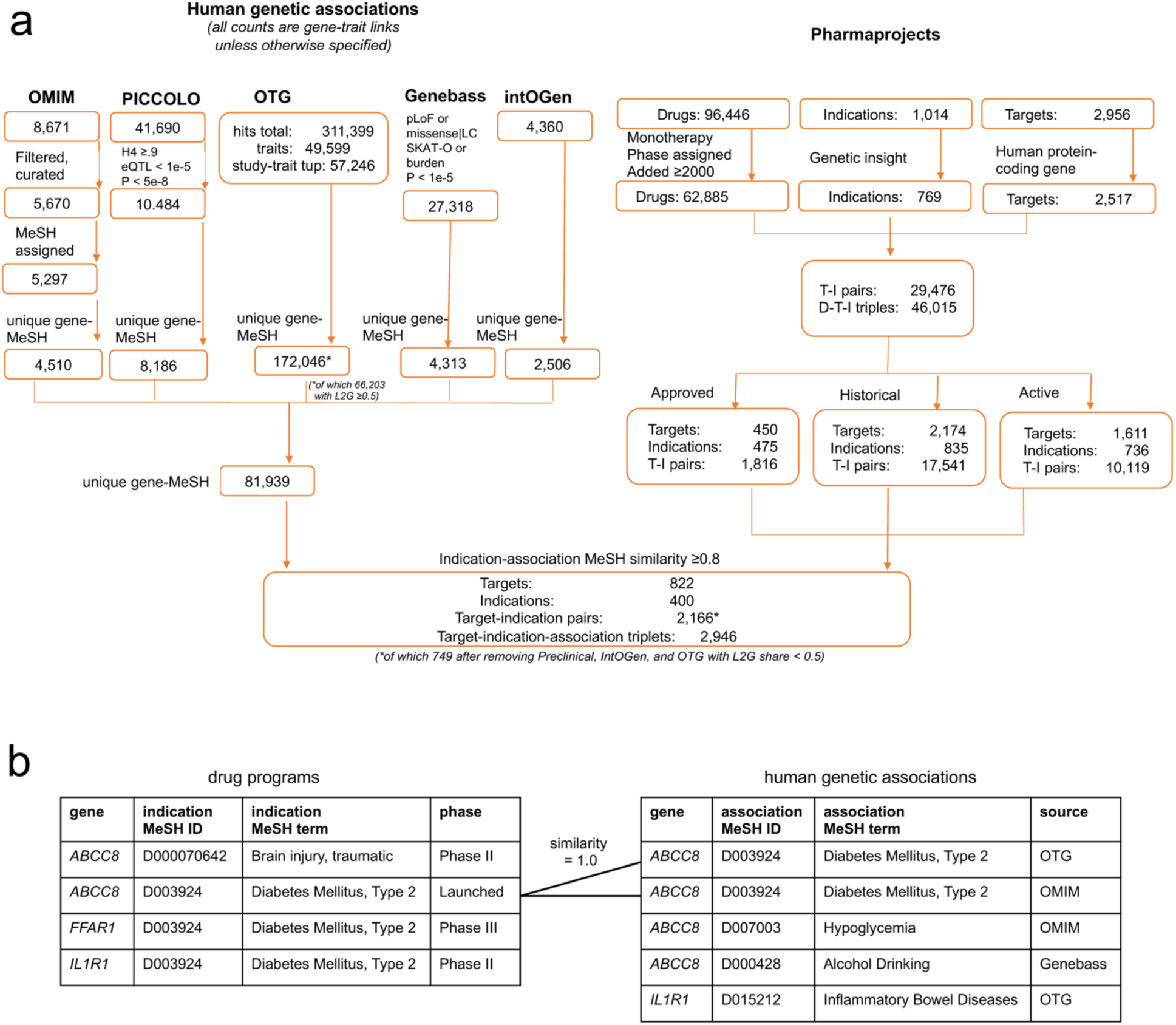
Data processing schematic. **A)** Dataset size, filters, and join process for Pharmaprojects and human genetic evidence. Note that a drug can be assigned multiple targets, and can be approved for multiple indications. The entire analysis described herein has also been run restricted to only those drugs with exactly one target annotated (Figures S1-S11). **B)** Illustration of the definition of genetic support. A table of drug development programs with one row per target-indication pair (left) is joined to a table of human genetic associations based on the identity of the gene encoding the drug target and the similarity between the drug indication MeSH term and the genetically associated trait MeSH term being ≥0.8. Drug program rows with a joined row in the genetic associations table are considered to have genetic support.

**Extended Data Figure 2.**
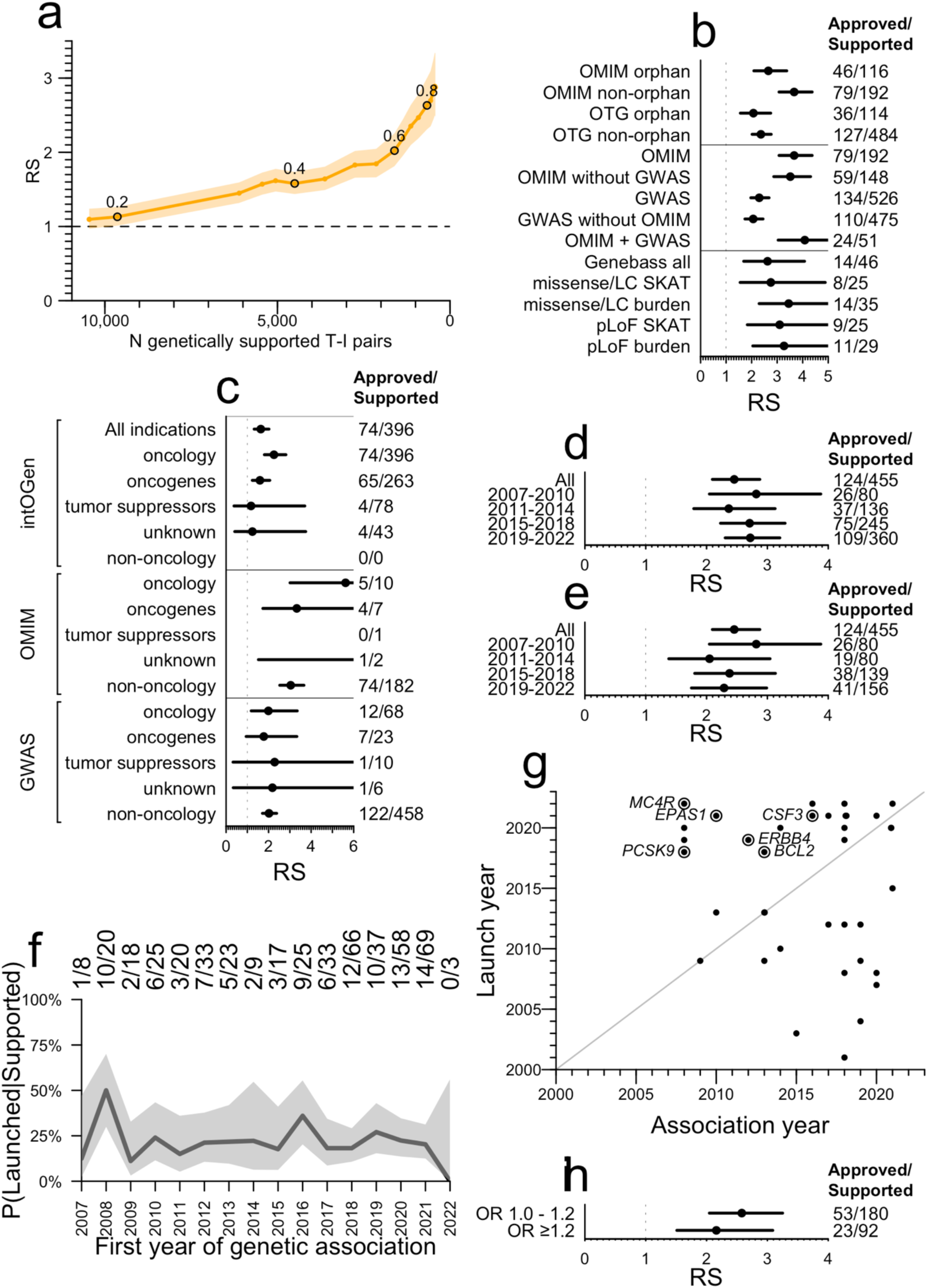
Further analysis of influence of characteristics of genetic associations on relative success. **A)** Sensitivity of RS to the similarity threshold between the MeSH ID for the genetically associated trait and the MeSH ID for the clinically developed indication. The threshold is varied by units of 0.05 (labels) and the results are plotted as RS (y axis) versus number of genetically supported T-I pairs (x axis). **B)** Breakdown of OTG and OMIM RS values by whether any drug for each T-I pair has had orphan status assigned. **C)** RS for somatic genetic evidence from IntOGen versus germline genetic evidence, for oncology and non-oncology indications. Note that the approved/supported proportions displayed for the top two rows are identical because all IntOGen genetic support is for oncology indications, yet the RS is different because the number of non-supported approved and non-supported clinical stage programs is different. In other words, in the “All indications” row, there is a Simpson’s paradox that diminishes the apparent RS of IntOGen — IntOGen support improves success rate (see 2^nd^ row) but also selects for oncology, an area with low baseline success rate (as shown in Extended Data Fig. 6A). **D)** As for top panel of Figure 1D, but without removing replications or OMIM-supported T-I pairs. **E)** As for top panel of Figure 1D, removing replications but not removing OMIM-supported T-I pairs. **F)** Proportion of T-I pairs supported by a GWAS Catalog association that are launched (versus phase I-III) as a function of the year of first genetic association. **G)** Launched T-I pairs genetically supported by OTG GWAS, shown by year of launch (y axis) and year of first genetic association (x axis). Gene symbols are labeled for first approvals of targets with at least 5 years between association and launch. Of 104 OTG- supported launched T-I pairs (Fig. 1D), year of drug launch was available for N=38 shown here, of which 18 (47%) acquired genetic support only in or after the year of launch. The true proportion of launched T-I whose GWAS support is retrospective may be larger if the T-I with a missing launch year are more often older drug approvals less well annotated in Pharmaprojects. **H)** Lack of impact of GWAS Catalog lead SNP odds ratio (OR) on RS when using the same OR breaks as used by King et al^15^.

**Extended Data Figure 3.**
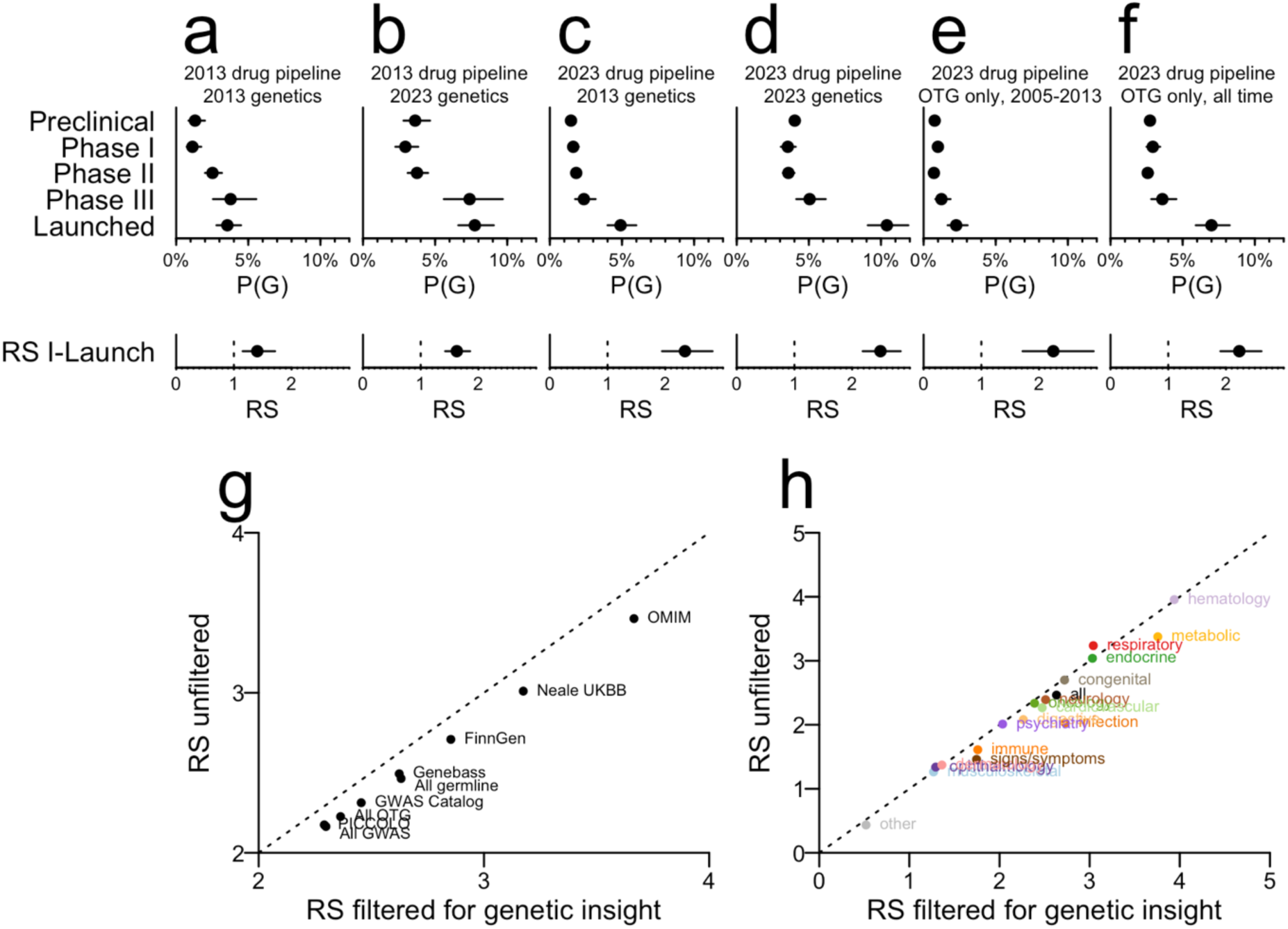
Sensitivity to changes in genetic data and drug pipeline over the past decade and to the ‘genetic insight’ filter. “2013” here indicates the data freezes from Nelson et al 2015^5^ (that study’s supplementary dataset 2 for genetics and supplementary dataset 3 for drug pipeline); “2023” indicates the data freezes in the present study. All datasets were processed using the current MeSH similarity matrix, and because “genetic insight” changes over time (more traits have been studied genetically now than in 2013), all panels are unfiltered for genetic insight (hence numbers in panel D differ from those in Fig. 1A). Every panel shows the proportion of combined (both historical and active) target-indication pairs with genetic support, or P(G), by development phase. **A)** 2013 drug pipeline and 2013 genetics. **B)** 2013 drug pipeline and 2023 genetics. **C)** 2023 drug pipeline and 2013 genetics. **D)** 2023 drug pipeline and 2023 genetics. **E)** 2023 drug pipeline with only OTG GWAS hits through 2013 and no other sources of genetic evidence. **F)** 2023 drug pipeline with only OTG GWAS hits for all years, no other sources of genetic evidence. We note that the increase in P(G) over the past decade^5^ is almost entirely attributable to new genetic evidence (e.g. contrast B vs. A, D vs. C, F vs. E) rather than changes in the drug pipeline (e.g. compare A vs. C, B vs. D). In contrast, the increase in RS is due mostly to changes in the drug pipeline (compare C, D, E, F vs. A, B), in line with theoretical expectations outlined by Hingorani et al^16^ and consistent with the findings of King et al^15^. We note that both the contrasts in this figure, and the fact that genetic support is so often retrospective (Extended Data Fig. 2G) suggest that P(G) will continue to rise in coming years. Because all panels here are unfiltered for genetic insight, we also show the difference in RS across **G)** sources of genetic evidence and **H)** therapy areas when this filter is removed. In general, removing this filter decreases RS by 0.17; this varies only slightly between sources and areas. The largest impact is seen in Infection, where removing the filter drops the RS from 2.73 to 2.03. The relatively minor impact of removing the genetic insight filter is consistent with the findings of King et al^15^, who varied the minimum number of genetic associations required for an indication to be included, and found that risk ratio for progression (i.e. RS) was slightly diminished when the threshold was reduced.

**Extended Data Figure 4.**
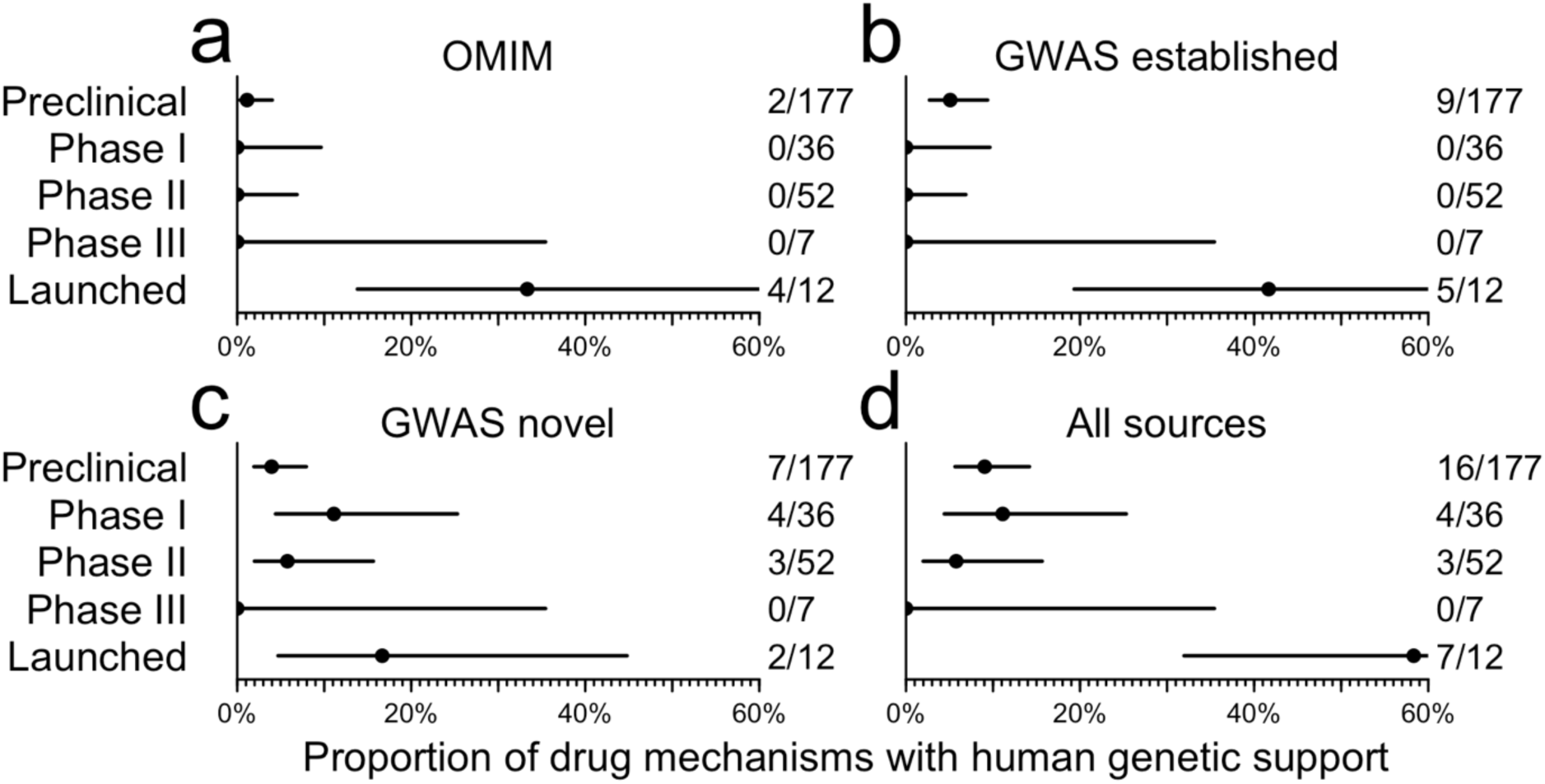
Proportion of type 2 diabetes drug targets with human genetic support by highest phase reached. A) OMIM, B) established (2019 and earlier) GWAS genes, C) novel (new in Vujkovic 2020 or Suzuki 2023) GWAS genes, or D) any of the above. See Methods for details on GWAS dataset processing.

**Extended Data Figure 5.**
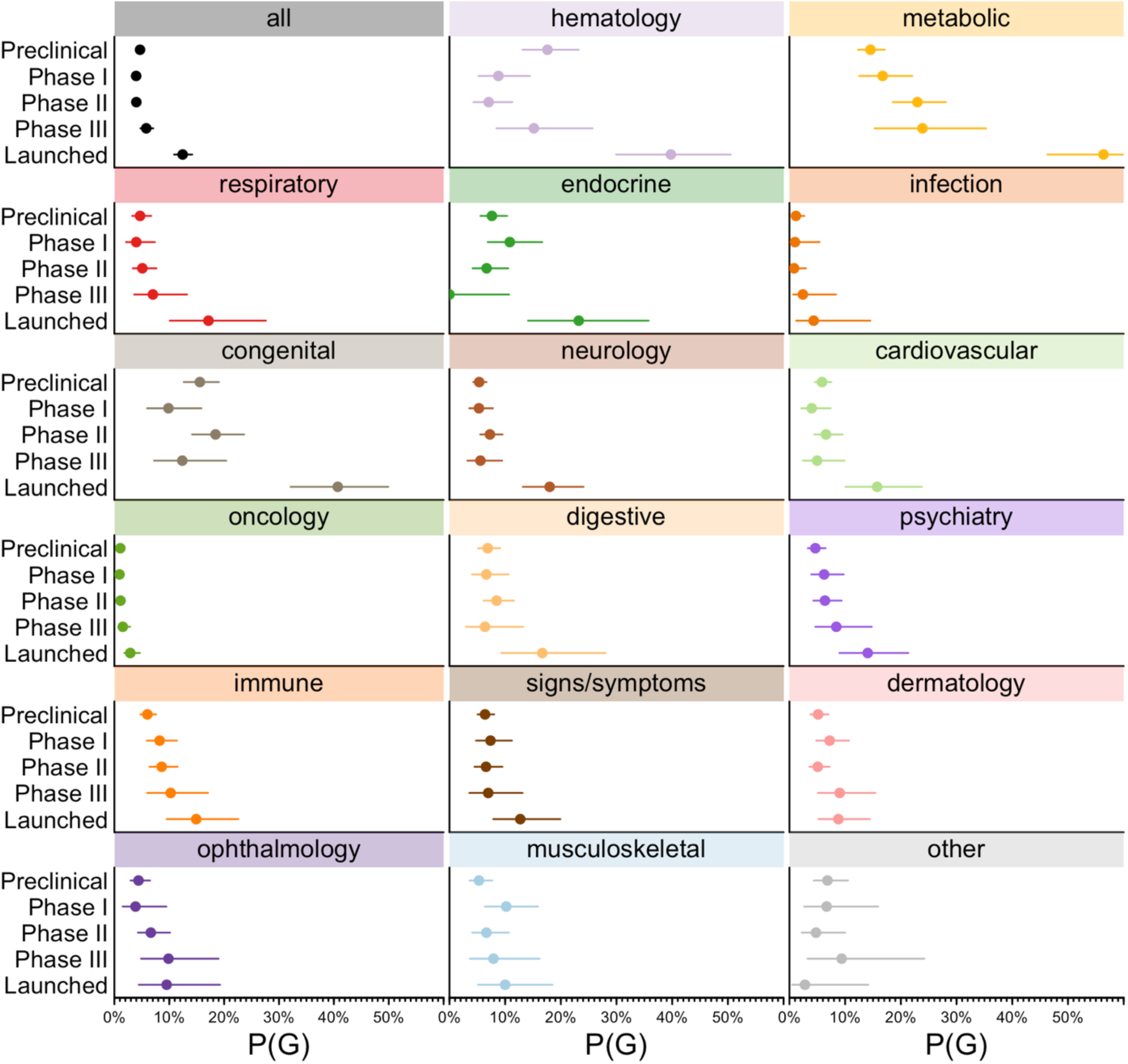
P(G) by phase versus therapy area. Each panel represents one therapy area, and shows the proportion of target-indication pairs in that area with genetic support, or P(G), by development phase.

**Extended Data Figure 6.**
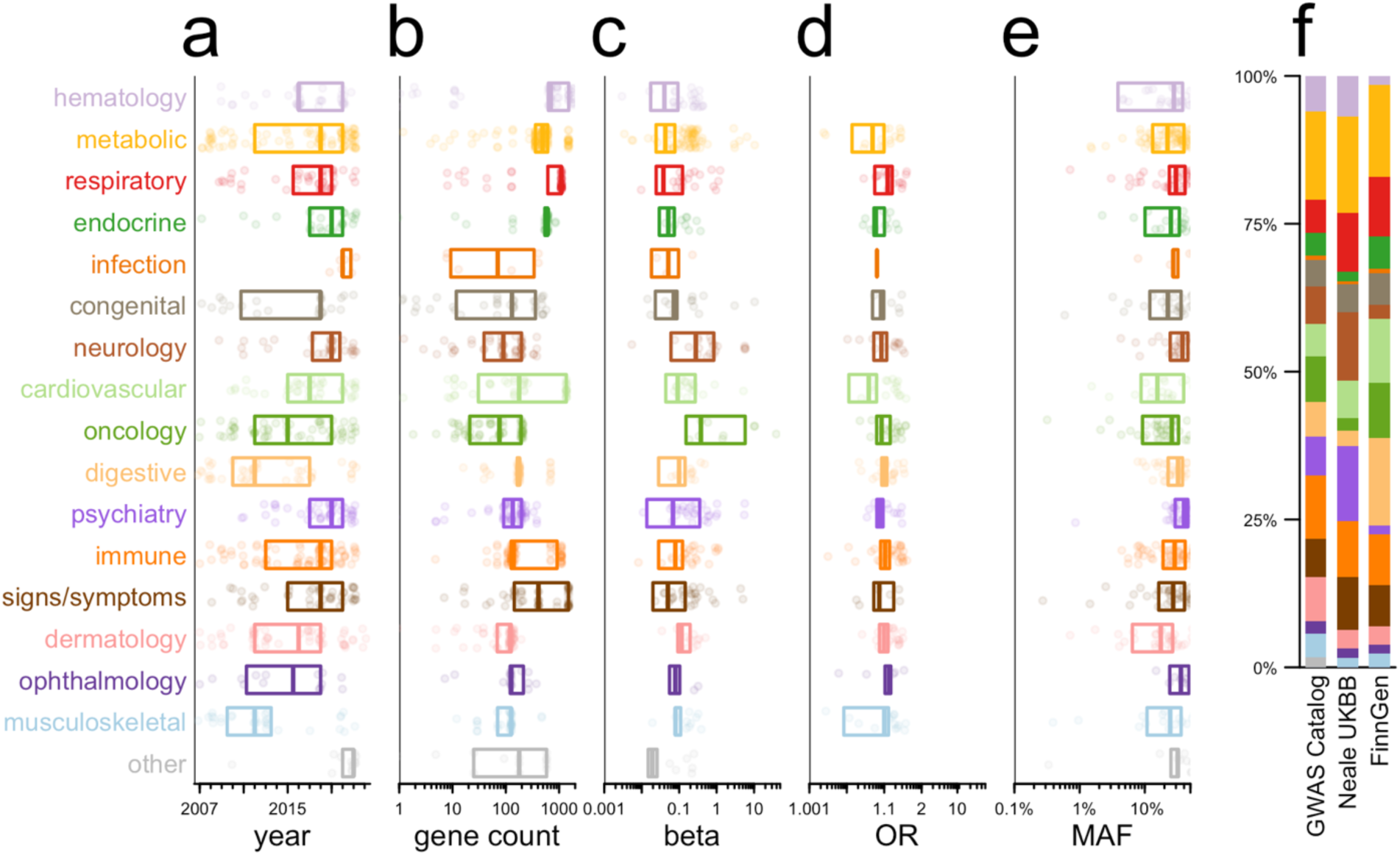
Confounding between therapy areas and properties of supporting genetic evidence. In panels A-E, each point represents one GWAS Catalog-supported T-I pair in phase I through launched, and boxes represent medians and interquartile ranges. Each panel A-E represents the cross-tabulation of therapy areas versus the properties examined in Fig. 1D. Kruskal-Wallis tests treat each variable as continuous, while chi-squared tests are applied to the discrete bins used in Figure 1D. **A**) Year of discovery, Kruskal-Wallis P = 1.1e-11, chi-squared P = 2.9e-16; **B**) gene count, Kruskal-Wallis P = 6.2e-35, chi-squared P = 7.1e-47; **C**) beta, Kruskal-Wallis P = 1.2e-5, chi-squared P = 1.7e-7; **D**) absolute odds ratio, Kruskal-Wallis P = 2.5e-5, chi-squared P = 4.3e-6 **E**) minor allele frequency, Kruskal-Wallis P = 5.7e-4, chi-squared P = 4.3e-3 **F**) Barplot of therapy areas of genetically supported T-I by source of GWAS data within OTG, chi-squared P = 2.4e-7.

**Extended Data Figure 7.**
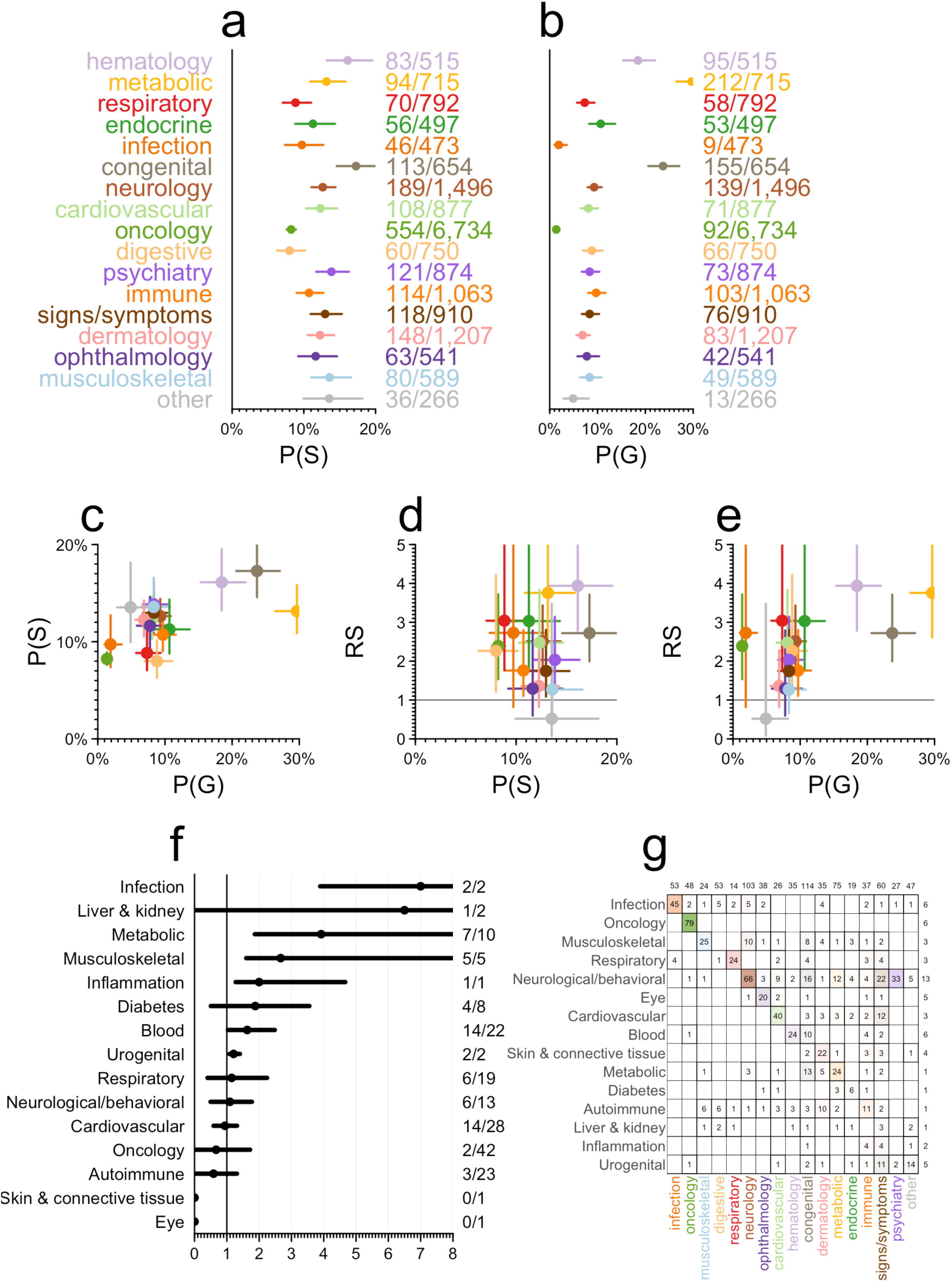
Further analyses of differences in relative success among therapy areas. **A)** Probability of success, P(S), by therapy area, with Wilson 95% confidence intervals. Fractions at right show the number of launched T-I pairs (numerator) and number of T-I pairs reaching at least phase I (denominator). **B)** Probability of genetic support, P(G), by therapy area, with Wilson 95% confidence intervals. Fractions at right show the number of genetically supported T-I pairs reaching at least phase I (numerator) and total number of T-I pairs reaching at least phase I (denominator). **C)** P(S) vs. P(G), **D)** RS s. P(S), and **E)** RS vs. P(G) across therapy areas, with crosshairs representing 95% confidence intervals on both dimensions. **F)** Re-analysis of RS (x axis) broken down by therapy area using data from supplementary table 6 of Nelson et al. 2015^5^. **G)** Confusion matrix showing the categorization of unique drug indications into therapy areas in Nelson et al 2015 versus current. Note that the current categorization is based on each indication’s position in the MeSH ontological tree and one indication can appear in >1 area, see Methods for details. Marginals along the top and right sides indicate the number of indication MeSH IDs in each drug pipeline dataset that are not present in the other drug pipeline dataset.

**Extended Data Figure 8.**
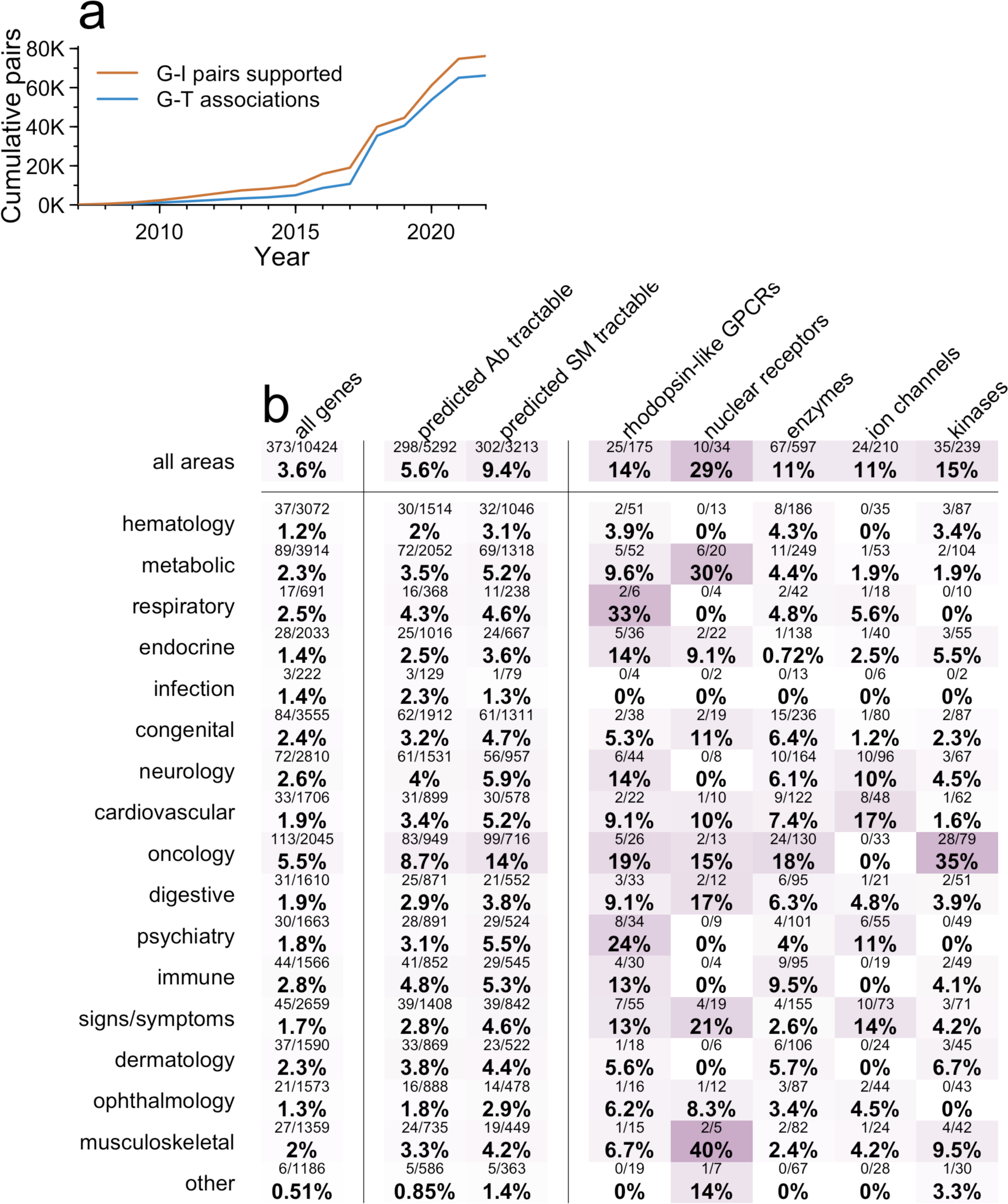
Level of utilization of genetic support among targets. As for Figure 3, but grouped by target instead of T-I pair. Thus, the denominator for each cell is the number of targets with at least one genetically supported indication, and each target counts towards the numerator if at least one genetically supported indication has reached phase I.

***Figures S1-S11. The three main and eight extended data figures restricted to drugs with one target only.***

## Notes

### Competing Interest Statement

MRN and CLD are employees of Deerfield. MRN is an employee of Genscience. EVM and JP are consultants to Deerfield. EVM also acknowledges speaking fees from Eli Lilly, consulting fees from Alnylam, and research support, unrelated to the current work, from Ionis, Sangamo, and Gate.

### Funding Statement

This study did not receive any funding

### Summary of Updates

This manuscript has been extensively revised to improve clarity and address feedback. Changes include updating of OMIM and IntOGen data to latest available as well as additional analyses and figures to better describe the study, data, and results. None of the changes had a material impact on the results and conclusions.

## REFERENCES

1. DiMasi JA, Grabowski HG, Hansen RW. Innovation in the pharmaceutical industry: New estimates of R&D costs. J Health Econ. 2016 May;47:20–33. PMID: 26928437

2. Hay M, Thomas DW, Craighead JL, Economides C, Rosenthal J. Clinical development success rates for investigational drugs. Nat Biotechnol. 2014 Jan;32(1):40–51. PMID: 24406927

3. Wong CH, Siah KW, Lo AW. Estimation of clinical trial success rates and related parameters. Biostatistics. 2019 01;20(2):273–286. PMCID: PMC6409418

4. Thomas D, Chancellor D, Micklus A, LaFever S, Hay M, Chaudhuri S, Bowden R, Lo AW. Clinical Development Success Rates and Contributing Factors 2011–2020 [Internet]. 2021 p. 34. Available from: https://go.bio.org/rs/490-EHZ-999/images/ClinicalDevelopmentSuccessRates2011_2020.pdf

5. Nelson MR, Tipney H, Painter JL, Shen J, Nicoletti P, Shen Y, Floratos A, Sham PC, Li MJ, Wang J, Cardon LR, Whittaker JC, Sanseau P. The support of human genetic evidence for approved drug indications. Nat Genet. 2015 Aug;47(8):856–860. PMID: 26121088

6. Diogo D, Tian C, Franklin CS, Alanne-Kinnunen M, March M, Spencer CCA, Vangjeli C, Weale ME, Mattsson H, Kilpeläinen E, Sleiman PMA, Reilly DF, McElwee J, Maranville JC, Chatterjee AK, Bhandari A, Nguyen KDH, Estrada K, Reeve MP, Hutz J, Bing N, John S, MacArthur DG, Salomaa V, Ripatti S, Hakonarson H, Daly MJ, Palotie A, Hinds DA, Donnelly P, Fox CS, Day-Williams AG, Plenge RM, Runz H. Phenome-wide association studies across large population cohorts support drug target validation. Nat Commun. 2018 16;16(1):4285. PMCID: PMC6191429

7. Plenge RM, Scolnick EM, Altshuler D. Validating therapeutic targets through human genetics. Nat Rev Drug Discov. 2013 Aug;12(8):581–594. PMID: 23868113

8. Musunuru K, Kathiresan S. Genetics of Common, Complex Coronary Artery Disease. Cell. 2019 Mar 21;177(1):132–145. PMID: 30901535

9. Trajanoska K, Bhérer C, Taliun D, Zhou S, Richards JB, Mooser V. From target discovery to clinical drug development with human genetics. Nature. 2023 Aug;620(7975):737–745. PMID: 37612393

10. Burgess S, Mason AM, Grant AJ, Slob EAW, Gkatzionis A, Zuber V, Patel A, Tian H, Liu C, Haynes WG, Hovingh GK, Knudsen LB, Whittaker JC, Gill D. Using genetic association data to guide drug discovery and development: Review of methods and applications. Am J Hum Genet. 2023 Feb 2;110(2):195–214. PMCID: PMC9943784

11. Carss KJ, Deaton AM, Del Rio-Espinola A, Diogo D, Fielden M, Kulkarni DA, Moggs J, Newham P, Nelson MR, Sistare FD, Ward LD, Yuan J. Using human genetics to improve safety assessment of therapeutics. Nat Rev Drug Discov. 2023 Feb;22(2):145–162. PMID: 36261593

12. Nguyen PA, Born DA, Deaton AM, Nioi P, Ward LD. Phenotypes associated with genes encoding drug targets are predictive of clinical trial side effects. Nat Commun. 2019 05;05(1):1579. PMCID: PMC6450952

13. Eric Vallabh Minikel, Matthew R Nelson. Human genetic evidence enriched for side effects of approved drugs. medRxiv. 2023 Dec 13;2023.12.12.23299869.

14. Visscher PM, Brown MA, McCarthy MI, Yang J. Five years of GWAS discovery. Am J Hum Genet. 2012 Jan 13;90(1):7–24. PMCID: PMC3257326

15. King EA, Davis JW, Degner JF. Are drug targets with genetic support twice as likely to be approved? Revised estimates of the impact of genetic support for drug mechanisms on the probability of drug approval. PLoS Genet. 2019;15(12):e1008489. PMCID: PMC6907751

16. Hingorani AD, Kuan V, Finan C, Kruger FA, Gaulton A, Chopade S, Sofat R, MacAllister RJ, Overington JP, Hemingway H, Denaxas S, Prieto D, Casas JP. Improving the odds of drug development success through human genomics: modelling study. Sci Rep. 2019 Dec 11;9(1):18911. PMCID: PMC6906499

17. Reay WR, Cairns MJ. Advancing the use of genome-wide association studies for drug repurposing. Nat Rev Genet. 2021 Oct;22(10):658–671. PMID: 34302145

18. Vujkovic M, Keaton JM, Lynch JA, Miller DR, Zhou J, Tcheandjieu C, Huffman JE, Assimes TL, Lorenz K, Zhu X, Hilliard AT, Judy RL, Huang J, Lee KM, Klarin D, Pyarajan S, Danesh J, Melander O, Rasheed A, Mallick NH, Hameed S, Qureshi IH, Afzal MN, Malik U, Jalal A, Abbas S, Sheng X, Gao L, Kaestner KH, Susztak K, Sun YV, DuVall SL, Cho K, Lee JS, Gaziano JM, Phillips LS, Meigs JB, Reaven PD, Wilson PW, Edwards TL, Rader DJ, Damrauer SM, O’Donnell CJ, Tsao PS, HPAP Consortium, Regeneron Genetics Center, VA Million Veteran Program, Chang KM, Voight BF, Saleheen D. Discovery of 318 new risk loci for type 2 diabetes and related vascular outcomes among 1.4 million participants in a multi-ancestry meta-analysis. Nat Genet. 2020 Jun 15; PMID: 32541925

19. Suzuki K, Hatzikotoulas K, Southam L, Taylor HJ, Yin X, Lorenz KM, Mandla R, Huerta-Chagoya A, Rayner NW, Bocher O, Ana Luiza de SVA, Sonehara K, Namba S, Lee SSK, Preuss MH, Petty LE, Schroeder P, Vanderwerff B, Kals M, Bragg F, Lin K, Guo X, Zhang W, Yao J, Kim YJ, Graff M, Takeuchi F, Nano J, Lamri A, Nakatochi M, Moon S, Scott RA, Cook JP, Lee JJ, Pan I, Taliun D, Parra EJ, Chai JF, Bielak LF, Tabara Y, Hai Y, Thorleifsson G, Grarup N, Sofer T, Wuttke M, Sarnowski C, Gieger C, Nousome D, Trompet S, Kwak SH, Long J, Sun M, Tong L, Chen WM, Nongmaithem SS, Noordam R, Lim VJY, Tam CHT, Joo YY, Chen CH, Raffield LM, Prins BP, Nicolas A, Yanek LR, Chen G, Brody JA, Kabagambe E, An P, Xiang AH, Choi HS, Cade BE, Tan J, Alaine Broadaway K, Williamson A, Kamali Z, Cui J, Adair LS, Adeyemo A, Aguilar-Salinas CA, Ahluwalia TS, Anand SS, Bertoni A, Bork-Jensen J, Brandslund I, Buchanan TA, Burant CF, Butterworth AS, Canouil M, Chan JCN, Chang LC, Chee ML, Chen J, Chen SH, Chen YT, Chen Z, Chuang LM, Cushman M, Danesh J, Das SK, Janaka de Silva H, Dedoussis G, Dimitrov L, Doumatey AP, Du S, Duan Q, Eckardt KU, Emery LS, Evans DS, Evans MK, Fischer K, Floyd JS, Ford I, Franco OH, Frayling TM, Freedman BI, Genter P, Gerstein HC, Giedraitis V, González-Villalpando C, González-Villalpando ME, Gordon-Larsen P, Gross M, Guare LA, Hackinger S, Han S, Hattersley AT, Herder C, Horikoshi M, Howard AG, Hsueh W, Huang M, Huang W, Hung YJ, Hwang MY, Hwu CM, Ichihara S, Ikram MA, Ingelsson M, Islam MT, Isono M, Jang HM, Jasmine F, Jiang G, Jonas JB, Jørgensen T, Kandeel FR, Kasturiratne A, Katsuya T, Kaur V, Kawaguchi T, Keaton JM, Kho AN, Khor CC, Kibriya MG, Kim DH, Kronenberg F, Kuusisto J, Läll K, Lange LA, Lee KM, Lee MS, Lee NR, Leong A, Li L, Li Y, Li-Gao R, Lithgart S, Lindgren CM, Linneberg A, Liu CT, Liu J, Locke AE, Louie T, Luan J, Luk AO, Luo X, Lv J, Lynch JA, Lyssenko V, Maeda S, Mamakou V, Mansuri SR, Matsuda K, Meitinger T, Metspalu A, Mo H, Morris AD, Nadler JL, Nalls MA, Nayak U, Ntalla I, Okada Y, Orozco L, Patel SR, Patil S, Pei P, Pereira MA, Peters A, Pirie FJ, Polikowsky HG, Porneala B, Prasad G, Rasmussen-Torvik LJ, Reiner AP, Roden M, Rohde R, Roll K, Sabanayagam C, Sandow K, Sankareswaran A, Sattar N, Schönherr S, Shahriar M, Shen B, Shi J, Shin DM, Shojima N, Smith JA, So WY, Stančáková A, Steinthorsdottir V, Stilp AM, Strauch K, Taylor KD, Thorand B, Thorsteinsdottir U, Tomlinson B, Tran TC, Tsai FJ, Tuomilehto J, Tusie-Luna T, Udler MS, Valladares-Salgado A, van Dam RM, van Klinken JB, Varma R, Wacher-Rodarte N, Wheeler E, Wickremasinghe AR, van Dijk KW, Witte DR, Yajnik CS, Yamamoto K, Yamamoto K, Yoon K, Yu C, Yuan JM, Yusuf S, Zawistowski M, Zhang L, Zheng W, VA Million Veteran Program, AMED GRIFIN Diabetes Initiative Japan, Project BJ, BioBank PM, Center RG, Consortium eMERGE, International Consortium for Blood Pressure (ICBP), Meta-Analyses of Glucose and Insulin-Related Traits Consortium (MAGIC), Raffel LJ, Igase M, Ipp E, Redline S, Cho YS, Lind L, Province MA, Fornage M, Hanis CL, Ingelsson E, Zonderman AB, Psaty BM, Wang YX, Rotimi CN, Becker DM, Matsuda F, Liu Y, Yokota M, Kardia SLR, Peyser PA, Pankow JS, Engert JC, Bonnefond A, Froguel P, Wilson JG, Sheu WHH, Wu JY, Geoffrey Hayes M, Ma RCW, Wong TY, Mook-Kanamori DO, Tuomi T, Chandak GR, Collins FS, Bharadwaj D, Paré G, Sale MM, Ahsan H, Motala AA, Shu XO, Park KS, Jukema JW, Cruz M, Chen YDI, Rich SS, McKean-Cowdin R, Grallert H, Cheng CY, Ghanbari M, Tai ES, Dupuis J, Kato N, Laakso M, Köttgen A, Koh WP, Bowden DW, Palmer CNA, Kooner JS, Kooperberg C, Liu S, North KE, Saleheen D, Hansen T, Pedersen O, Wareham NJ, Lee J, Kim BJ, Millwood IY, Walters RG, Stefansson K, Goodarzi MO, Mohlke KL, Langenberg C, Haiman CA, Loos RJF, Florez JC, Rader DJ, Ritchie MD, Zöllner S, Mägi R, Denny JC, Yamauchi T, Kadowaki T, Chambers JC, Ng MCY, Sim X, Below JE, Tsao PS, Chang KM, McCarthy MI, Meigs JB, Mahajan A, Spracklen CN, Mercader JM, Boehnke M, Rotter JI, Vujkovic M, Voight BF, Morris AP, Zeggini E. Multi-ancestry genome-wide study in >2.5 million individuals reveals heterogeneity in mechanistic pathways of type 2 diabetes and complications. medRxiv. 2023 Mar 31;2023.03.31.23287839. PMCID: PMC10081410

20. Lommatzsch M, Brusselle GG, Canonica GW, Jackson DJ, Nair P, Buhl R, Virchow JC. Disease-modifying anti-asthmatic drugs. Lancet. 2022 Apr 23;399(10335)1664–1668. PMID: 35461560

21. Mortberg MA, Vallabh SM, Minikel EV. Disease stages and therapeutic hypotheses in two decades of neurodegenerative disease clinical trials. Sci Rep. 2022 Oct 21;12(1):17708. PMCID: PMC9587287

22. Minikel EV, Karczewski KJ, Martin HC, Cummings BB, Whiffin N, Rhodes D, Alföldi J, Trembath RC, van Heel DA, Daly MJ, Genome Aggregation Database Production Team, Genome Aggregation Database Consortium, Schreiber SL, MacArthur DG. Evaluating drug targets through human loss-of-function genetic variation. Nature. 2020 May;581(7809):459–464. PMCID: PMC7272226

23. Ference BA, Ginsberg HN, Graham I, Ray KK, Packard CJ, Bruckert E, Hegele RA, Krauss RM, Raal FJ, Schunkert H, Watts GF, Borén J, Fazio S, Horton JD, Masana L, Nicholls SJ, Nordestgaard BG, van de Sluis B, Taskinen MR, Tokgözoglu L, Landmesser U, Laufs U, Wiklund O, Stock JK, Chapman MJ, Catapano AL. Low-density lipoproteins cause atherosclerotic cardiovascular disease. 1. Evidence from genetic, epidemiologic, and clinical studies. A consensus statement from the European Atherosclerosis Society Consensus Panel. Eur Heart J. 2017 Aug 21;38(32):2459–2472. PMCID: PMC5837225

24. Scannell JW, Bosley J, Hickman JA, Dawson GR, Truebel H, Ferreira GS, Richards D, Treherne JM. Predictive validity in drug discovery: what it is, why it matters and how to improve it. Nat Rev Drug Discov. 2022 Dec;21(12):915–931. PMID: 36195754

25. Sun BB, Kurki MI, Foley CN, Mechakra A, Chen CY, Marshall E, Wilk JB, Biogen Biobank Team, Chahine M, Chevalier P, Christé G, FinnGen, Palotie A, Daly MJ, Runz H. Genetic associations of protein-coding variants in human disease. Nature. 2022 Mar;603(7899):95–102. PMCID: PMC8891017

26. Citeline Pharmaprojects [Internet]. [cited 2023 Aug 30]. Available from: https://web.archive.org/web/20230830135309/https://www.citeline.com/en/products-services/clinical/pharmaprojects

27. Painter JL. Toward automating an inference model on unstructured terminologies: OXMIS case study. Adv Exp Med Biol. 2010;680:645–651. PMID: 20865550

28. Mountjoy E, Schmidt EM, Carmona M, Schwartzentruber J, Peat G, Miranda A, Fumis L, Hayhurst J, Buniello A, Karim MA, Wright D, Hercules A, Papa E, Fauman EB, Barrett JC, Todd JA, Ochoa D, Dunham I, Ghoussaini M. An open approach to systematically prioritize causal variants and genes at all published human GWAS trait-associated loci. Nat Genet. 2021 Nov;53(11):1527–1533. PMCID: PMC7611956

29. Zheng J, Haberland V, Baird D, Walker V, Haycock PC, Hurle MR, Gutteridge A, Erola P, Liu Y, Luo S, Robinson J, Richardson TG, Staley JR, Elsworth B, Burgess S, Sun BB, Danesh J, Runz H, Maranville JC, Martin HM, Yarmolinsky J, Laurin C, Holmes MV, Liu JZ, Estrada K, Santos R, McCarthy L, Waterworth D, Nelson MR, Smith GD, Butterworth AS, Hemani G, Scott RA, Gaunt TR. Phenome-wide Mendelian randomization mapping the influence of the plasma proteome on complex diseases. Nat Genet. 2020 Oct;52(10):1122–1131. PMCID: PMC7610464

30. Sollis E, Mosaku A, Abid A, Buniello A, Cerezo M, Gil L, Groza T, Güneş O, Hall P, Hayhurst J, Ibrahim A, Ji Y, John S, Lewis E, MacArthur JAL, McMahon A, Osumi-Sutherland D, Panoutsopoulou K, Pendlington Z, Ramachandran S, Stefancsik R, Stewart J, Whetzel P, Wilson R, Hindorff L, Cunningham F, Lambert SA, Inouye M, Parkinson H, Harris LW. The NHGRI-EBI GWAS Catalog: knowledgebase and deposition resource. Nucleic Acids Res. 2023 Jan 6;51(D1):D977–D985. PMCID: PMC9825413

31. Kurki MI, Karjalainen J, Palta P, Sipilä TP, Kristiansson K, Donner KM, Reeve MP, Laivuori H, Aavikko M, Kaunisto MA, Loukola A, Lahtela E, Mattsson H, Laiho P, Della Briotta Parolo P, Lehisto AA, Kanai M, Mars N, Rämö J, Kiiskinen T, Heyne HO, Veerapen K, Rüeger S, Lemmelä S, Zhou W, Ruotsalainen S, Pärn K, Hiekkalinna T, Koskelainen S, Paajanen T, Llorens V, Gracia-Tabuenca J, Siirtola H, Reis K, Elnahas AG, Sun B, Foley CN, Aalto-Setälä K, Alasoo K, Arvas M, Auro K, Biswas S, Bizaki-Vallaskangas A, Carpen O, Chen CY, Dada OA, Ding Z, Ehm MG, Eklund K, Färkkilä M, Finucane H, Ganna A, Ghazal A, Graham RR, Green EM, Hakanen A, Hautalahti M, Hedman ÅK, Hiltunen M, Hinttala R, Hovatta I, Hu X, Huertas-Vazquez A, Huilaja L, Hunkapiller J, Jacob H, Jensen JN, Joensuu H, John S, Julkunen V, Jung M, Junttila J, Kaarniranta K, Kähönen M, Kajanne R, Kallio L, Kälviäinen R, Kaprio J, FinnGen, Kerimov N, Kettunen J, Kilpeläinen E, Kilpi T, Klinger K, Kosma VM, Kuopio T, Kurra V, Laisk T, Laukkanen J, Lawless N, Liu A, Longerich S, Mägi R, Mäkelä J, Mäkitie A, Malarstig A, Mannermaa A, Maranville J, Matakidou A, Meretoja T, Mozaffari SV, Niemi MEK, Niemi M, Niiranen T, O Donnell CJ, Obeidat ME, Okafo G, Ollila HM, Palomäki A, Palotie T, Partanen J, Paul DS, Pelkonen M, Pendergrass RK, Petrovski S, Pitkäranta A, Platt A, Pulford D, Punkka E, Pussinen P, Raghavan N, Rahimov F, Rajpal D, Renaud NA, Riley-Gillis B, Rodosthenous R, Saarentaus E, Salminen A, Salminen E, Salomaa V, Schleutker J, Serpi R, Shen HY, Siegel R, Silander K, Siltanen S, Soini S, Soininen H, Sul JH, Tachmazidou I, Tasanen K, Tienari P, Toppila-Salmi S, Tukiainen T, Tuomi T, Turunen JA, Ulirsch JC, Vaura F, Virolainen P, Waring J, Waterworth D, Yang R, Nelis M, Reigo A, Metspalu A, Milani L, Esko T, Fox C, Havulinna AS, Perola M, Ripatti S, Jalanko A, Laitinen T, Mäkelä TP, Plenge R, McCarthy M, Runz H, Daly MJ, Palotie A. FinnGen provides genetic insights from a well-phenotyped isolated population. Nature. 2023 Jan;613(7944):508–518. PMCID: PMC9849126

32. Guo C, Sieber KB, Esparza-Gordillo J, Hurle MR, Song K, Yeo AJ, Yerges-Armstrong LM, Johnson T, Nelson MR. Identification of putative effector genes across the GWAS Catalog using molecular quantitative trait loci from 68 tissues and cell types. bioRxiv. 2019 Jan 1;808444.

33. Karczewski KJ, Solomonson M, Chao KR, Goodrich JK, Tiao G, Lu W, Riley-Gillis BM, Tsai EA, Kim HI, Zheng X, Rahimov F, Esmaeeli S, Grundstad AJ, Reppell M, Waring J, Jacob H, Sexton D, Bronson PG, Chen X, Hu X, Goldstein JI, King D, Vittal C, Poterba T, Palmer DS, Churchhouse C, Howrigan DP, Zhou W, Watts NA, Nguyen K, Nguyen H, Mason C, Farnham C, Tolonen C, Gauthier LD, Gupta N, MacArthur DG, Rehm HL, Seed C, Philippakis AA, Daly MJ, Davis JW, Runz H, Miller MR, Neale BM. Systematic single-variant and gene-based association testing of thousands of phenotypes in 394,841 UK Biobank exomes. Cell Genomics. 2022 Sep 14;2(9):100168.

34. Lin D. An information-theoretic definition of similarity. Icml. 1998. p. 296–304.

35. Resnik P. Semantic similarity in a taxonomy: An information-based measure and its application to problems of ambiguity in natural language. Journal of artificial intelligence research. 1999;11:95–130.

36. The UniProt Consortium. UniProt: the universal protein knowledgebase. Nucleic Acids Res. 2017 04;45(D1):D158–D169. PMCID: PMC5210571

37. Ochoa D, Hercules A, Carmona M, Suveges D, Baker J, Malangone C, Lopez I, Miranda A, Cruz-Castillo C, Fumis L, Bernal-Llinares M, Tsukanov K, Cornu H, Tsirigos K, Razuvayevskaya O, Buniello A, Schwartzentruber J, Karim M, Ariano B, Martinez Osorio RE, Ferrer J, Ge X, Machlitt-Northen S, Gonzalez-Uriarte A, Saha S, Tirunagari S, Mehta C, Roldán-Romero JM, Horswell S, Young S, Ghoussaini M, Hulcoop DG, Dunham I, McDonagh EM. The next-generation Open Targets Platform: reimagined, redesigned, rebuilt. Nucleic Acids Res. 2023 Jan 6;51(D1):D1353–D1359. PMCID: PMC9825572

